# The Metabolomic Signature of Stressful Life Events

**DOI:** 10.64898/2026.04.02.26350045

**Authors:** Yumeng Tian, Ruifang Li-Gao, Tahani Alshehri, Christopher R. Brydges, Matthias Arnold, Siamak Mahmoudiandehkordi, Gabi Kastenmüller, Dennis O. Mook-Kanamori, Frits R. Rosendaal, Erik J. Giltay, Lin Xu, Jiao Wang, Rick Jansen, Thomaz Bastiaanssen, Brenda W.J.H. Penninx, Rima Kaddurah-Daouk, Yuri Milaneschi

**Affiliations:** Department of Clinical Epidemiology, Leiden University Medical Center, Leiden, the Netherlands; Biochemistry Department, College of Science, King Saud University, Riyadh, Saudi Arabi; West Coast Metabolomics Center, University of California, Davis, CA, United States; Institute of Computational Biology, Helmholtz Zentrum München-German Research Center for Environmental Health, Neuherberg, Germany; Department of Psychiatry and Behavioral Sciences, Duke University, Durham, NC, USA; Department of Psychiatry, Leiden University Medical Center, Leiden, the Netherlands; School of Public Health, Sun Yat-Sen University, Guangzhou, China; School of Public Health, The University of Hong Kong, Hong Kong, China; Greater Bay Area Public Health Research Collaboration, Guangdong-Hong Kong-Macao, China; Department of Psychiatry, Amsterdam UMC, Vrije Universiteit Amsterdam, Amsterdam, The Netherlands; Amsterdam Public Health, Mental Health program, Amsterdam, The Netherlands; Amsterdam Neuroscience, Mood, Anxiety, Psychosis, Sleep & Stress program, Amsterdam, The Netherlands; Department of Medicine, Duke University, Durham, NC, USA; Duke Institute of Brain Sciences, Duke University, Durham, NC, USA

**Keywords:** Stressful life events, Metabolome, Metabolomics, Lipid metabolism, Fatty acid, Bile acid

## Abstract

Stressful life events impact individual’s functionality and contribute to disease outcomes, yet the biological pathways underlying life stress remain unclear. We characterized the metabolomic profiles of stressful life events using data from 3,264 participants (5,163 observations) of the Dutch NESDA cohort. 98 metabolites were identified, with upregulated metabolites overrepresented in phosphatidylethanolamine and downregulated metabolites overrepresented in fatty acid metabolism. 92 of these metabolites were available in the Dutch NEO cohort (N=599): 11 were significantly replicated including six lipids (e.g., three bile acids (glycochenodeoxycholate 3-sulfate)), one carbohydrate, and one xenobiotic. 21 overlapping metabolites were additionally available in the Chinese GBCS cohort (N=200): 10-undecenoate (11:1n1) (fatty acid) and glycochenodeoxycholate 3-sulfate (bile acid) showed consistent associations across both Dutch and Chinese cohorts. Stressful life events are associated with metabolic dysregulation, particularly involving fatty acid and bile acid pathways, highlighting promising biological targets to reduce the impact of stress on mental and somatic health.

## Introduction

Stressful life events, such as the death of a loved one, job loss, divorce, or serious illness, are significant disruptions that may occur throughout the life course. These events can severely impact a person’s functionality and well-being and contribute to adverse health outcomes^1^. Evidence from the past decades revealed that individuals experiencing severe stressful life events had an increased susceptibility to somatic disorders, particularly cardiometabolic diseases^2–4^. The underlying mechanisms linking stressful life events to disease outcomes could be explained by the dysregulation in mood, behavior and physiology^5^. Specifically, affective dysregulations may manifest as elevated levels of depression or anxiety^6,7^. Behavioral changes (e.g., poor diet quality, excessive smoking and alcohol consumption) may occur as the coping response to stressful life events^8^. Physiological disruptions may involve neuroendocrine and autonomic response systems, such as activation of the hypothalamic-pituitary-adrenal (HPA) axis, contributing to hemodynamic alterations, endothelial dysfunction, immune dysregulation and inflammation^9,10^. These mechanisms are to known to alter circulating metabolite profiles which may represent sensitive indicators of the disrupted balance between environmental stressors and physiological regulation. As suggested by previous systematic reviews, metabolic disruption may play a mediating role in the association between life stress (encompassing both past and present stressors, as well as various disadvantages) and disease outcomes, for instance cardiometabolic diseases^10,11^. Therefore, exploring the metabolomic characterization of stressful life events may provide insights into the mechanisms underlying stressful life events-triggered disease outcomes and inform preventive strategies.

Individuals with stressful life events have an increased cardiometabolic risk^12,13^. To elucidate the metabolite alterations underlying this risk, previous studies employing targeted metabolomics platforms reported associations of stressful life events with insulin- and lipid-related pathways, including fasting glucose^14,15^, proinsulin c-peptide^16^ or insulin^17^, triglycerides^18^, apolipoprotein B^19^, and steroid hormones^20^. However, most of these studies were limited in the number and classes of metabolites assessed. Untargeted metabolomics can capture a broader spectrum of the metabolome, thereby enabling the discovery of novel pathobiological mechanisms linking stressful life events to disease outcomes. Metabolomic analysis of stressful life events in a larger population with such extended coverage of the metabolome is warranted.

In the current study, we aimed to comprehensively investigate the metabolomic signature of stressful life events using data from the Netherlands Study of Depression and Anxiety (NESDA) cohort including >5,000 observations with assessment on stressful life events and 820 metabolites from the untargeted Metabolon HD4 platform. External replication analyses for the significantly associated metabolites were conducted in two independent cohorts, the Netherlands Epidemiology of Obesity (NEO) study and the Chinese Guangzhou Biobank Cohort Study (GBCS), further testing the robustness of the main findings across populations with diverse ancestral and cultural contexts, lifestyles, and disease profiles.

## Methods

### Discovery analysis in the NESDA

#### Study design

The workflow of the current study is shown in Figure 1. NESDA, used as the discovery cohort, is an ongoing longitudinal study on the course and consequences of depression^21^. Briefly, at baseline 2,329 individuals with a remitted or current Diagnostic and Statistical Manual of Mental Disorders Fourth Edition (DSM-IV) based depressive (major depressive disorder, dysthymia) and/or anxiety disorder (panic disorder, social phobia, agoraphobia, generalized anxiety disorder) and 652 healthy controls, yielding a total of 2,981 participants were included from the community, primary care, and secondary care settings between 2004 and 2007. Follow-up visits were taken place after 1, 2, 4, 6, and 9 years. Additionally, the sibling sub-study involving 367 participants was recruited at the 9-year follow-up. Each assessment included a diagnostic interview to assess the presence of depressive and anxiety disorders, a medical examination, and several questionnaires on symptom severity, other clinical characteristics and lifestyle. Fasting blood samples were collected at baseline, 6-year follow-up in the original NESDA cohort and in the sibling sub-study. The study was approved by the ethical review boards of participating centers, and all participants provided informed consent. The present study used 5,163 observations from 3,264 participants with available stressful life events and metabolomics measurements: 2,804 from baseline and 2,008 at 6-year follow-up (of which 1,901 overlapped with baseline), and 351 from the sibling sub-study.

**Figure 1.**
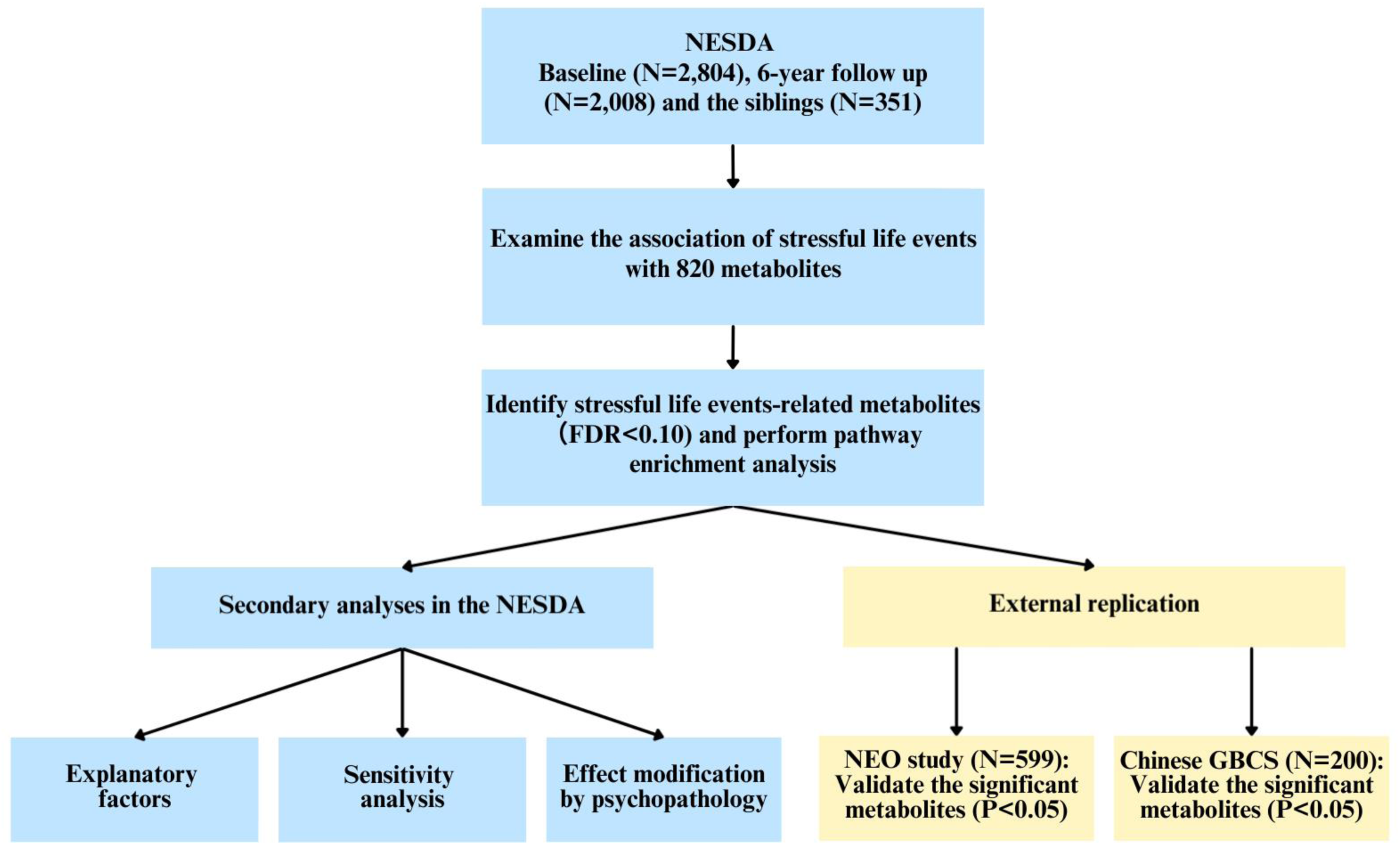
Workflow of the current study. Notes: NESDA: Netherlands Study of Depression and Anxiety; NEO: Netherlands Epidemiology of Obesity; GBCS: Guangzhou Biobank Cohort Study; FDR: false discovery rate.

#### Assessment of stressful life events

We derived the stressful life events sum scores from the List of Threatening Events (LTE) Questionnaire^22^, which assesses twelve major stressful life events over the past year using 12 dichotomous (yes/no) items, including events such as good friend or close relative died and facing serious financial problems. The LTE focuses on event occurrence rather than subjective appraisal of events. Detailed items of LTE questionnaire are shown in the Supplementary Appendix. Scores range from 0 to 12, reflecting the total number of events experienced. This scale has high validity and reliability^22^. Participants were assessed at baseline, 6-year follow-up and the sibling sub-study of NESDA. The stressful life events scores were scaled to standard deviation (SD) units for comparison.

#### Measurements of metabolomics and quality control

Metabolomic profiles were measured using the Metabolon^TM^ Discovery HD4 platform (Metabolon Inc., Durham, North Carolina, USA)^21^. The assessment and quality control of metabolite levels was good, and has been reported in the previous NESDA studies^23^. Briefly, fasting plasma samples were stored at −80 °C and sent for analyses in two batches. Missing metabolite values were imputed with the k-nearest neighbor approach (k=10). The dataset was log2 transformed and values were scaled to SD units for comparison. Finally, 820 metabolites, including 681 characterized (metabolites with known identity) and 139 uncharacterized (metabolites with unknown identity), were measured in the NESDA^23^. Detailed information is shown in the Supplementary Appendix.

#### Covariates

Sex (men/women), age (continuous), education level (basic/intermediate/high), shipment batch (if applicable), and wave (if applicable) were included as covariates in the main analysis (model 1). In an additional model, we examined the impact of further adjustment for lifestyle and health-related factors that may potentially represents mechanisms explaining the associations detected (model 2). Lifestyle factors included smoking (never/former/current), alcohol consumption (continuous), and total physical activity level (continuous). Health-related factors included BMI (continuous), self-reported chronic diseases score (based on 20 conditions) (continuous), and lipid-lowering medication use (no/yes). Measurements of these factors are described in the Supplementary Appendix.

#### Statistical Analysis

Characteristics of the study population in NESDA was expressed as a mean with SD or percentages. In the main analyses, we used linear mixed-effects models (R lme4 and lmerTest packages) to examine the associations between the total number of stressful life events and metabolite levels assessed at baseline, the sub-sibling study and 6-year follow-up, thereby utilizing full statistical power by including data available at multiple waves. The models were specified with a Gaussian distribution, two random intercepts at the family and participant levels to account for within-family and within-subject correlations and were estimated via maximum likelihood. Analyses were adjusted for sex, age, education level, shipment batch, and wave (model 1). To correct for multiple testing across the metabolites, the false discovery rate (FDR) correction was applied to the model, and metabolites with an FDR-corrected P value <0.10 were carried forward in subsequent analyses. Pathway enrichment analyses for the metabolites significantly positively or negatively associated with stressful life events were performed using pathways pre-assigned by Metabolon^23^, including 10 metabolite classes (super-pathways) and 95 pathways (sub-pathways). Enrichment analyses^23^ were based on the Fisher exact test.

Furthermore, we performed three sets of secondary analyses. 1) To study the mechanisms potentially explaining the significant associations detected, we additionally adjusted the models for lifestyle and health-related factors, including BMI, alcohol consumption, smoking, total physical activity level, self-reported chronic diseases score (based on 20 conditions), and lipid-lowering medication use (model 2). 2) To examine the impact on the detected associations of lipid-lowering medication use, particularly relevant for lipoproteins and fatty acids^24^, we performed sensitivity analysis excluding observations in which the participants reported using these medications identified by the Anatomical Therapeutic Chemical (ATC) code C10 (the use of medication was assessed based on drug container inspection of all medication used in the past month). 3) Since NESDA is enriched of individuals with depression and anxiety, and both conditions have specific metabolomic signatures^23,25^ and were significantly related to stressful life events^6,7^, we further examined whether the stressful life events-metabolites associations detected differed between participants with psychopathology (depressive and/or anxiety disorders) and healthy controls. This effect modification analysis was performed by adding the interaction term of stressful life events with psychopathology status to the linear mixed-effects models. Psychopathology was defined by the presence of a DSM-IV lifetime diagnoses of depressive (major depressive disorder or dysthymia) and/or anxiety disorders (panic disorder, social phobia, generalized anxiety disorder, or agoraphobia) assessed using Composite International Diagnostic Interview (CIDI version 2.1)^26^.

#### External replications in the NEO study and GBCS

The NEO study is a population-based cohort study of individuals aged between 45 and 65 years with a self-reported body mass index (BMI) of 27 kg/m^2^ or higher, living in the greater area of Leiden (in the West of the Netherlands)^27^. In addition, all inhabitants aged between 45 and 65 years from one municipality (Leiderdorp) were invited, irrespective of their BMI. Recruitment of participants started in September 2008 and completed at the end of September 2012. A subset of participants (n=599) from the Leiderdorp with data on stressful life events and fasting blood with metabolomics was utilized for the replication analyses^28^. Participants in the NEO study were assessed with the same LTE questionnaire applied in NESDA^22^. Then, stressful life events scores were scaled to SD units for comparison. Metabolomic profiles were measured at baseline using the same Metabolon^TM^ Discovery HD4 platform applied in NESDA^28^. The quality control has been reported in the previous NEO study, which adopted a specific imputation workflow for missing values using the multivariate imputation by chained equations (MICE) approach for metabolomic data^29^. Then, the dataset was log2 transformed, and values were scaled to SD units for comparison.

The GBCS is a four-way collaboration among Guangzhou Twelfth People’s Hospital and the Universities of Hong Kong, Birmingham and Sun Yat-Sen^30^. Participants aged 50 years or above were enrolled in Guangzhou, the capital city of Guangdong Province in Southern China, from September 2003 to January 2008. All participants were invited to return for the first (March 2008 to December 2012) follow-up examinations. The follow-up questionnaire and clinical and laboratory examinations were largely similar to those conducted at baseline. A subset of participants (n=200) with measurements of stressful life events and fasting blood with metabolomics was utilized for replication analyses. Detailed information is shown in Supplementary Appendix. The stressful life events sum scores were assessed at baseline using ten dichotomous items (i.e., yes/no), which evaluate the presence or absence of ten major life events over the past year, with total scores ranging from 0 to 10^30^. These ten items are comparable to those in the LTE questionnaire, and also assess life events such as unemployment, major illness, and death of a close family member. Detailed information of these ten items is shown in the Supplementary Appendix. Previous GBCS study used these ten items to assess life events and examined its association with depression, reporting a positive association with the risk of depression^31^. These results are in line with findings from prior studies using the LTE questionnaire to assess stressful life events^6,32^. The stressful life events scores were scaled to SD units for comparison in GBCS. Metabolomic profiles were measured at the first follow-up using the untargeted liquid chromatography-mass spectrometry (LC-MS)-based metabolomics platform (Level One 500)^33^. This platform could detect more than 2,000 metabolites in accordance with the Metabolomics Standards Initiative and report relative (semi-quantitative) metabolite abundances. Metabolite values below the limit of detection were imputed with half of the minimal values. Then, the dataset was log2 transformed, and values were scaled to SD units for comparison. Although the measurements of stressful life events and metabolites in the GBCS differed from those applied in NESDA, we aimed to replicate the findings in a Chinese population to identify consistent associations across diverse ancestral and cultural contexts, lifestyles, and disease profiles. Detailed information is shown in the Supplementary Appendix.

The associations between stressful life events and metabolites identified in NESDA were tested in replication cohorts using linear regressions adjusted for sex, age and education level. Metabolites available in NESDA were mapped to NEO and GBCS using Chemical, Human Metabolome Database (HMDB), or PUBCHEM IDs. Replication was deemed for metabolites with estimate directionally concordant with NESDA and statistically significant at nominal P <0.05, two sided. Finally, association estimates for metabolites available across all three cohorts were pooled with inverse-variance-weighted (IVW) random-effects meta-analyses.

## Results

### Main analyses

Table 1 shows the characteristics of participants at baseline (n=2,804), the siblings (n=351) and participants at 6-year follow-up (n=2,008) in the NESDA. The mean age of participants at baseline and the siblings was 42.1 and 51.2 years, respectively, with 66.2% and 55.6% women. Around half of the participants in the baseline (51.3%) and in the sibling (50.1%) samples reported at least one stressful life event over the past year. At 6-year follow-up 74.3% of the participants reported at least one stressful life event.

**Table 1.** Characteristics of baseline, the siblings and 6-year follow-up in the NESDA.

| Characteristics | Baseline | Siblings | 6-year follow-up |
| --- | --- | --- | --- |
| Number of participants | 2,804 | 351 | 2,008 |
| Age, years, mean (SD) | 42.1 (13.0) | 51.2 (13.1) | 48.2 (13.1) |
| Sex, woman, n (%) | 1,857 (66.2) | 195 (55.6) | 1,317 (65.6) |
| BMI, mean (SD) | 25.5 (5.0) | 25.8 (4.9) | 24.9 (7.8) |
| Education level, n (%) |  |  |  |
| Basic | 186 (6.6) | 9 (2.6) | 94 (4.7) |
| Intermediate | 1,600 (57.1) | 178 (51.1) | 983 (49.0) |
| High | 1,018 (36.3) | 161 (46.3) | 931 (46.4) |
| Smoking status, n (%) |  |  |  |
| Never | 789 (28.1) | 122 (34.9) | 599 (29.9) |
| Former | 946 (33.7) | 143 (40.9) | 843 (42.0) |
| Current | 1,069 (38.1) | 85 (24.3) | 563 (28.1) |
| Alcohol consumption, drinks/week, mean (SD) | 7.1 (9.9) | 7.1 (8.9) | 6.1 (8.6) |
| Total physical activity, MET hours/week, mean (SD) | 61.5 (51.7) | 71.9 (61.5) | 65.9 (57.1) |
| Psychopathology status, n (%) |  |  |  |
| Healthy control | 626 (22.3) | 177 (50.4) | 470 (23.4) |
| Lifetime depression or anxiety | 2,178 (77.7) | 174 (49.6) | 1,538 (76.6) |
| Self-reported chronic diseases score (based on 20 conditions), mean (SD) | 0.89 (1.07) | 0.89 (1.06) | 0.79 (0.98) |
| Lipid-lowering drug use, yes, n (%) | 200 (7.1) | 49 (14.0) | 226 (11.3) |
| Stressful life events total score, mean (SD) | 0.88 (1.11) | 0.73 (0.89) | 1.49 (1.35) |
| Stressful life events, n (%) |  |  |  |
| 0 | 1,365 (48.7) | 175 (49.9) | 516 (25.7) |
| 1 | 800 (28.5) | 117 (33.3) | 640 (31.9) |
| 2 | 379 (13.5) | 43 (12.3) | 463 (23.1) |
| 3+ | 260 (9.3) | 16 (4.6) | 389 (19.4) |

BMI: body mass index; SD: standard deviation; NESDA: Netherlands Study of Depression and Anxiety.

The abundance of, in total, 820 blood metabolites, covering ten super-pathways of metabolites (e.g., lipids (38%), amino acids (21%) and xenobiotics (11%)) and 95 sub-pathways, were determined and tested for their association with stressful life events (Supplementary Table 1). Totally, stressful life events were associated with 98 metabolites (FDR<0.10) after adjustment for sex, age, education level, shipment batch, and wave (model 1) (Figure 2). Of these, 69 stressful life events-related metabolites showed increased levels, whereas 29 showed decreased levels. Nearly half of these metabolites were lipids (46%), followed by amino acids (17%) and xenobiotics (9%). The top 10 metabolites were one xenobiotic (i.e., o-cresol sulfate), four amino acids-related metabolites (i.e., 5-hydroxylysine, 5-methylthioadenosine (MTA), gamma-carboxyglutamate and tyramine O-sulfate), four lipids (i.e., tetradecadienoate (14:2), 1-oleoyl-GPE (18:1), 1-linoleoyl-GPE (18:2) and 3,4-dihydroxybutyrate), and one partially characterized molecule (i.e., branched-chain, straight-chain, or cyclopropyl 12:1 fatty acid).

**Figure 2.**
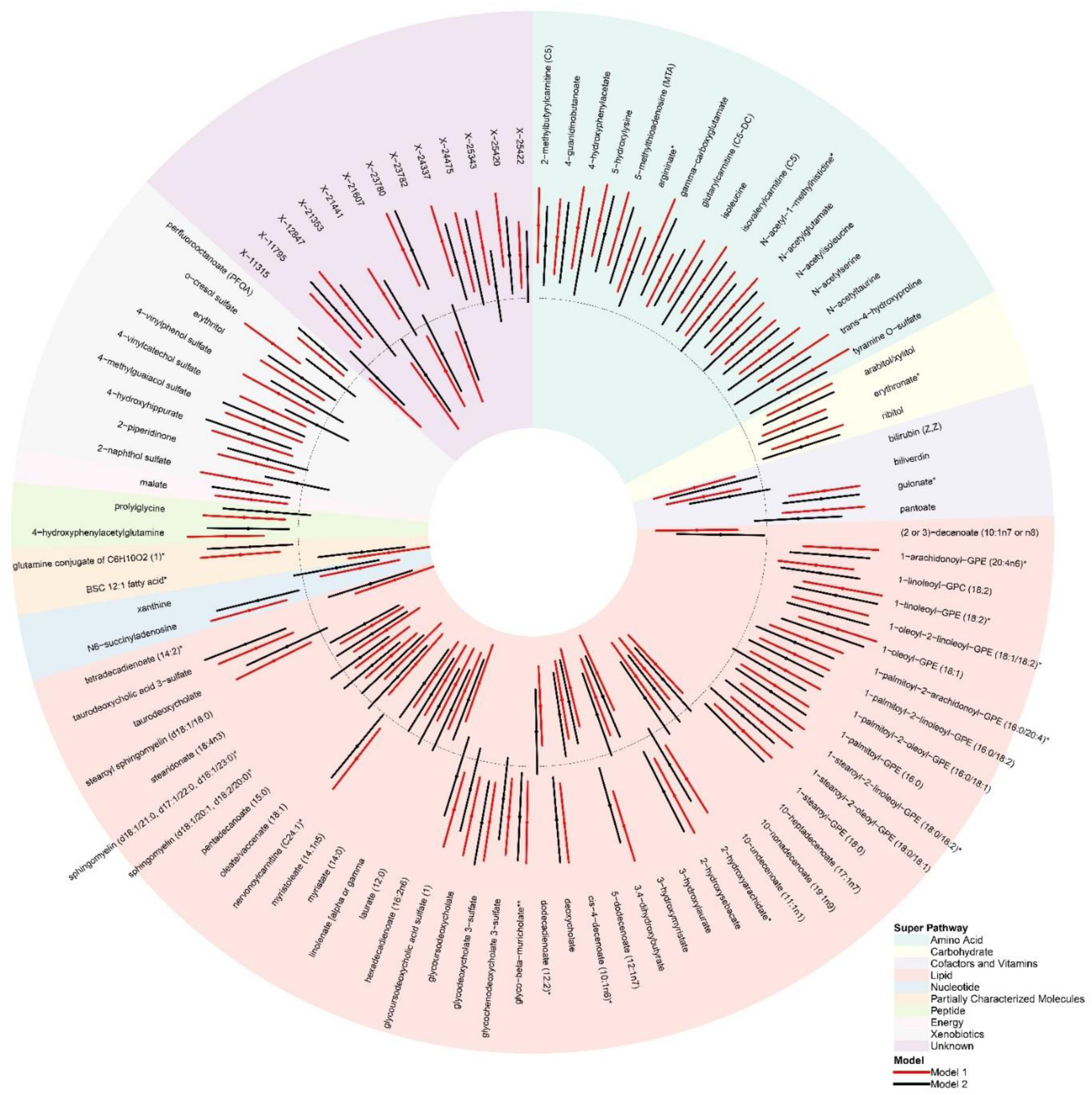
The association of stressful life events with 98 metabolites identified in the NESDA. Notes: (1) The dotted circle indicates the reference (standardized beta=0), and background color indicates the super-pathway of each metabolite. Metabolites positively associated with stressful life events are shown in the dotted outer circle, while negatively associated metabolites are shown in the dotted inner circle. (2) The significant associations of stressful life events with 98 metabolites were identified in model 1 (red line) based on the FDR<0.10. Of them, 69 stressful life events-related metabolites were higher, while 29 stressful life events-related metabolites were lower. (3) After additional adjustment in model 2 (black line), 34 stressful life event-related metabolites were non-significant (P≥0.05). (4) Model 1 was adjusted for sex, age, education level, wave and shipment batch. Model 2 was additionally adjusted for BMI, alcohol consumption, smoking, level of total physical activity, lipid-lowering drug use, and self-reported chronic diseases score (based on 20 conditions). (5) BMI: body mass index; FDR: false discovery rate; BSC 12:1 fatty acid*: branched-chain, straight-chain, or cyclopropyl 12:1 fatty acid*.

Using the 98 stressful life events-related metabolites, we performed pathway enrichment analyses for 85 metabolites (60 higher and 25 lower); 13 metabolites with uncharacterized chemical identity were excluded. The comprehensive results for the super- and sub-pathway enrichment analyses are shown in Supplementary Tables 2 and 3. Among the 10 super-pathways, negatively associated metabolites were significantly enriched in lipids (P=1.22×10^-3^, FDR=1.10×10^-2^, Supplementary Table 2). Considering the 95 sub-pathways, positively associated metabolites were overrepresented in the phosphatidylethanolamine sub-pathway (P=9.14×10^-4^, FDR=8.59×10^-2^), while negatively associated metabolites were overrepresented in medium-chain fatty acid (P=9.67×10^-5^, FDR=9.09×10^-3^) and long-chain monounsaturated fatty acid (MUFA) sub-pathways (P=8.52×10^-4^, FDR=4.00×10^-2^), respectively (Table 2).

**Table 2.** Pathway enrichment analysis of metabolites positively (n = 60) or negatively (n = 25) associated with stressful life events in the NESDA.

| Pathway | Positively associated metabolites (n=60) |  |  |  | Negatively associated metabolites (n=25) |  |  |
| --- | --- | --- | --- | --- | --- | --- | --- |
|  | Pathway size, n (%)<sup>a</sup> | P | FDR | Overlap, n (%)<sup>b</sup> | P | FDR | Overlap, n (%)<sup>b</sup> |
| Phosphatidylethanolamine (PE) | 10 (1.5) | 9.14e-04 | 8.59e-02 | 6 (10.0) | NS | NS | 0 |
| Medium chain fatty acid | 8 (1.2) | NS | NS | 0 | 9.67e-05 | 9.09e-03 | 5 (20.0) |
| Long-chain monounsaturated fatty acid | 7 (1.0) | NS | NS | 0 | 8.52e-04 | 4.00e-02 | 4 (16.0) |

<sup>a</sup>: Pathway size is the percentage of all metabolites (n = 681) assigned to the pathway.
<sup>b</sup>: Overlap is the number of metabolites (%) that are both in the pathway and in the significantly higher or lower metabolite set.
A total of 85 stressful life events-related metabolites (excluding 13 metabolites with uncharacterized chemical identity) were identified based on the FDR<0.10.
n: number of metabolites; FDR: false discovery rate.

### Secondary analyses

We examined the potential role of lifestyle and health-related factors in the 98 significant stressful life events-metabolite associations, by additionally adjusting the statistical model for lifestyle and health-related factors, including BMI, smoking, alcohol consumption, total physical activity level, the self-reported chronic diseases score (based on 20 conditions), and lipid-lowering medication use (model 2, results in Supplementary Table 4 and Figure 2). Generally, all 98 metabolites in model 2 showed effect size estimates in same directions as in model 1 (r=0.96, Supplementary Figure 1), although with a median reduction of 10.44% (Supplementary Table 4), suggesting that lifestyle and health-related factors partially explained the association between stressful life events and metabolites. Three xenobiotics, namely 2-naphthol sulfate, 4-vinylcatechol sulfate and o-cresol sulfate, showed the largest reduction in estimates (from 77.79% to 92.59%) and were no longer significantly associated with stressful life events. In contrast, 67 metabolites remained significantly associated with stressful life events in model 2, including 14 with decreased levels (e.g., 5-dodecenoate (12:1n7), a medium chain fatty acid) and 53 with increased levels (e.g., 1-oleoyl-2-linoleoyl-GPE (18:1/18:2), a phosphatidylethanolamine).

Furthermore, to examine the impact of lipid-lowering medication use on the 98 stressful life events-metabolite association we re-ran the analyses after excluding 377 participants using these medications (618 observations). Of the 98 associations, 97 metabolites remained statistically significant (Supplementary Table 5), and all effect estimates showed highly consistent effect with those from the full sample (r=0.99, Supplementary Figure 2) suggesting a minimal impact of lipid-lowering medication on the observed associations.

Finally, no significant interaction between stressful life events and psychopathology status was found for almost all 98 metabolites, with the only exception of 4-methylguaiacol sulfate (P for interaction=8.33×10^-4^, stratified analyses show that participants with lifetime depression or anxiety (β=0.053, P=3.98×10^-4^); healthy control (β=-0.068, P=3.73×10^-2^), Supplementary Table 6).

### External replications in the NEO study and GBCS

Supplementary Table 7 shows the baseline characteristics of the two replication cohorts, the NEO study (n=599) and the GBCS (n=200), indicating that the mean age was 55.8 and 63.4 years with 52.6% and 85.5% women, respectively. A total of 40.6% of participants in the NEO study and 7.1% of participants in the GBCS reported experiencing stressful life events over the past year. Of the 98 stressful life events-related metabolites identified in NESDA, 92 metabolites were available in the NEO study. The effect estimates for these 92 stressful life events-associated metabolites were moderately but significantly correlated between the two cohorts (r=0.40, P=1.60×10^-4^, Supplementary Figure 3), suggesting a general consistency in effect sizes. Specifically, 11 metabolites in the NEO study showed a consistent direction of association and were significantly associated (P<0.05) with stressful life events as illustrated in Figure 3. The 11 metabolites included six lipids (i.e., 1-palmitoyl-2-arachidonoyl-GPE (16:0/20:4), 1-palmitoyl-GPE (16:0), 3,4-dihydroxybutyrate, glycodeoxycholate 3-sulfate, glycochenodeoxycholate 3-sulfate and taurodeoxycholic acid 3-sulfate), one carbohydrate (i.e., ribitol), one xenobiotic (i.e., 2-naphthol sulfate) and three metabolites with uncharacterized chemical identity. In the GBCS, of the 98 stressful life events-related metabolites, 21 metabolites were available. The overall correlation between effect estimates in NESDA and GBCS was not significant (r=-0.30, P=0.25). None of the metabolites with directionally consistent effect estimates reached statistically significant in the GBCS (Supplementary Table 9). For 10-undecenoate (11:1n1), a metabolite in the medium chain fatty acid sub-pathway, the association approached threshold for statistical significance (β=-0.038, P=6.15×10^-2^). 20 metabolites were present in all three cohorts. As depicted in Figure 3, two of these were lipids, namely glycochenodeoxycholate 3-sulfate and 10-undecenoate (11:1n1), and consistently showed the same direction of association with stressful life events, although effect estimates in GBCS had wider variance likely due to relatively small sample size. Meta-analyses of estimates for 20 metabolites across the three cohorts yielded pooled effect estimates of β=0.056 (P=2.61×10^-2^) for glycochenodeoxycholate 3-sulfate and β=-0.037 (P=3.70×10^-3^) for 10-undecenoate (11:1n1) (Supplementary Table 10).

**Figure 3.**
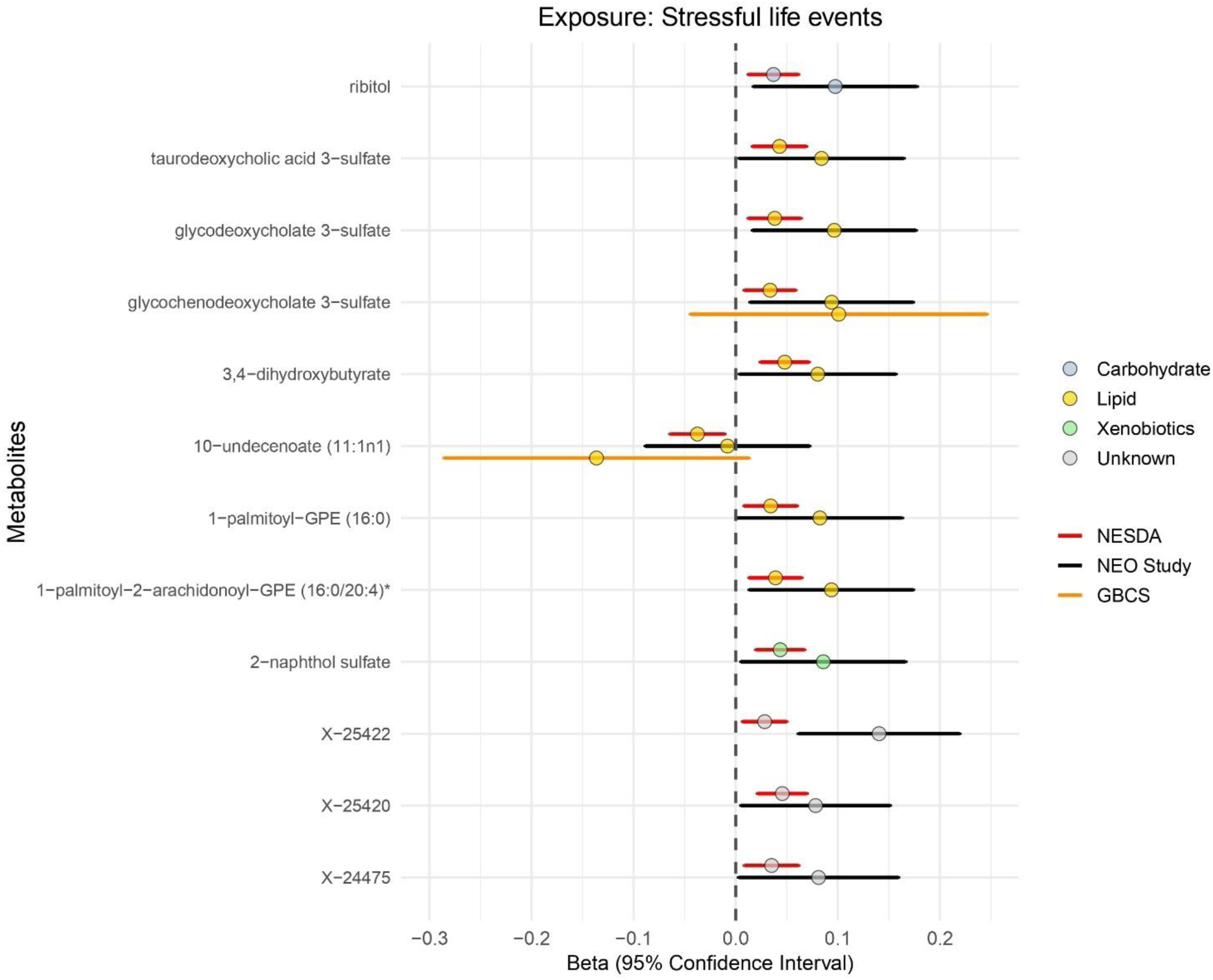
Significant replication of stressful life events-related metabolites in the NEO study and GBCS. Notes: (1) The dotted line indicates the reference (standardized beta=0) and the color in the circle indicates the class (super-pathway) of each metabolite. Positive beta indicates the higher metabolite levels found with stressful life events, while negative beta indicates the lower metabolite levels. (2) 98 stressful life events-related metabolites identified in the NESDA (FDR<0.10) were used for external replication. Of them, 92 metabolites could be matched in the NEO study using CHEM ID, and 21 metabolites could be matched in the GBCS using PUBCHEM and HMDB ID. (3) Detailed results were shown in the Supplementary Table 8, 9, and 10. Figure 3 summarized that 11 metabolites were significantly replicated (P<0.05) in the NEO study (black line), while 1 metabolite showed marginal significance (P=0.07) in the GBCS (yellow line). (4) In the NESDA, model 1 was adjusted for age, sex, education level, wave and shipment batch. In the NEO study and GBCS, model 1 was adjusted for age, sex and education level.

## Discussion

The present study is the largest to date systematically examining the association between recent stressful life events and a wide metabolic profile measured with an untargeted metabolomics platform. In the NESDA cohort including 5,163 observations from 3,264 participants, we identified 98 stressful life events-associated metabolites, of which 69 were positively associated and 29 were negatively associated. Characteristic patterns were observed, with positive associations of metabolites in the phosphatidylethanolamine pathway and negative associations in the medium-chain fatty acid and long-chain MUFA pathways. Notably, 11 stressful life events-related metabolites were nominally replicated in the NEO cohort, particularly three bile acids (i.e., glycodeoxycholate 3-sulfate, glycochenodeoxycholate 3-sulfate and taurodeoxycholic acid 3-sulfate). In the GBCS, one fatty acid (10-undecenoate (11:1n1)) replicated, approaching threshold for statistical significance. Among the 20 metabolites measured in all three cohorts, glycochenodeoxycholate 3-sulfate and 10-undecenoate (11:1n1) showed consistent associations with stressful life events across the Dutch cohorts (i.e., NESDA and NEO) and the Chinese GBCS cohort, suggesting the robustness of these metabolic alterations across populations with different cultural and ancestral backgrounds.

Stressful life events were associated with metabolites significantly enriched with alterations in lipid metabolism, including phosphatidylethanolamine metabolism. The elevated levels of six phosphatidylethanolamines, including 1-palmitoyl-2-arachidonoyl-GPE (16:0/20:4), were observed in individuals with stressful life events. This elevation was in line with previous studies, reporting significantly higher levels of phosphatidylethanolamines in the brain of mice exposed to unpredictable chronic stress^34^. Phosphatidylethanolamines, key phospholipids in lipoproteins, are particularly susceptible to oxidative modifications^35^, which contribute to inflammation and immune responses by accelerating the production of pro-inflammatory cytokines^36,37^. This function might link phosphatidylethanolamines (in particular those carrying oxidized fatty acids such as arachidonic acid (20:4)^38^) to disease pathogenesis, supported by evidence of elevated levels of 1-palmitoyl-2-arachidonoyl-GPE (16:0/20:4) in myocardial infarction patients^39^ and increased oxidized phosphatidylethanolamines in the plasma of individuals with aortic stenosis^40^ and the animal model of diabetes^41^.

Overall, the identified metabolic signature of stressful life events is highly convergent with that of cardiometabolic conditions, such as cardiovascular diseases and diabetes, potentially representing biological substrates connecting the onset of these conditions with stress-related dysregulations. Consistent with this interpretation, several stressful life events-related metabolites, including ribitol, 3,4-dihydroxybutyrate and 1-palmitoyl-GPE (16:0), might be involved in the link between stressful life events and cardiometabolic disease risks. For instance, a previous metabolomics study of coronary artery disease (CAD) involving 441 participants showed a positive association between higher levels of ribitol and CAD risk^42^. A similar association was observed with both microvascular and macrovascular progression^43^. All of those conditions are known risk factors for the development of cardiometabolic disease outcomes. Additionally, in previous metabolomics studies, 3,4-dihydroxybutyrate and 1-palmitoyl-GPE (16:0) were identified as potential biomarkers associated with diabetes complications^44,45^. Meanwhile, 1-palmitoyl-GPE (16:0) was also linked to subclinical coronary atherosclerosis through an increasing burden of noncalcified plaque^46^. Future studies are warranted to assess the potential mediating role of these metabolites in the association between stressful life events and cardiometabolic risks.

Different mechanisms may explain the association between stressful life events and metabolic dysregulation. Findings from analyses in NESDA suggested that lifestyle and somatic health factors might partially explain stressful life events-metabolite associations. Remarkably, stressful life events were associated with increased levels of xenobiotics, including o-cresol sulfate (i.e., the strongest association before adjusting for lifestyle factors), 2-naphthol sulfate and 4-vinylcatechol sulfate. However, such associations were substantially attenuated after additional adjustment for lifestyle and somatic health factors. These metabolites were identified as biomarkers of exposure to environmental pollutants generated by the incomplete combustion of organic materials, for instance, tobacco. Consistent with this interpretation, secondary analyses from a cohort of 1,082 Dutch participants examining the metabolomic profiles of several lifestyle factors demonstrated that smoking was associated with increased levels of 2-naphthol sulfate, o-cresol sulfate and 4-vinylcatechol sulfate^47^. Notably, in our study, other xenobiotics directly involved in tobacco metabolism, for instance cotinine, were excluded during quality control due to high missingness. The detection of three xenobiotics mentioned above suggests that these metabolite levels might be influenced by the combination of multiple exogenous exposures and endogenous metabolic processes. This notion is supported by a previous study of 602 American individuals showing that higher levels of xenobiotics, including o-cresol sulfate, were observed in patients with liver failure compared to those without disease outcome^48^. Therefore, the increased levels of xenobiotics observed in individuals experiencing stressful life events may reflect underlying lifestyle patterns, such as smoking, together with other co-occurring adverse somatic health conditions. These metabolites may, in turn, play a role in the development of cardiometabolic diseases^49^.

Nevertheless, the majority of stressful life events-metabolite associations remained largely independent from lifestyle and health-related factors, suggesting a more complex interplay between environmentally derived substrates and endogenous biological processes. For instance, 12 metabolites were involved in fatty acid metabolism, particularly the long-chain polyunsaturated fatty acid (PUFA) and long-chain MUFA pathways. These pathways also exhibited the enrichment of negatively associated metabolites. Interestingly, although these fatty acids are primarily derived from the diet, their biological processing is partly regulated by genetics^50^. Similarly, stearoyl-coA desaturase 1 (SCD1) gene clusters encode the rate-limiting enzymes SCD, directly regulating metabolic shifts towards production of long-chain MUFAs^51^. These desaturase-driven transforming collectively contributed to the synthesis of numerous individual long-chain PUFA and MUFA (e.g., myristoleate (14:1n5) and stearidonate (18:4n3)), which were potentially related to chronic inflammation^52^, cardiovascular health^53^, and neurological development^54^.

In discovery and replication analyses, we observed several stressful life events-related metabolites, including glycochenodeoxycholate 3-sulfate, glycodeoxycholate 3-sulfate and taurodeoxycholic acid 3-sulfate, three bile acids. 10-undecenoate (11:1n1) also showed a significant association with stressful life events. These metabolites have previously been linked to disruptions in gut microbiota composition^55,56^. For instance, a large study of 8,583 Swedish individuals examined the association of gut microbial species with metabolomics, indicating that certain species were linked to metabolite levels, particularly those involved in the secondary bile acid metabolism pathway and their precursors^55^. Notably, glycodeoxycholate 3-sulfate exhibited a strong positive correlation with *Anaerotruncus colihominis*, and such a pattern could also be observed for taurodeoxycholic acid 3-sulfate. Meanwhile, 10-undecenoate (11:1n1) had a positive correlation with *Veillonella rogosae*. Consistently, another study found that 10-undecenoate (11:1n1) and metabolites belonging to bile acid metabolism were associated with gut microbiota and predicted the gut microbial α-diversity^56^. In addition to these endogenous metabolites, three smoking-related xenobiotics, namely o-cresol sulfate, 2-naphthol sulfate and 4-vinylcatechol sulfate, have also been increasingly linked to gut microbiome via their precursors (i.e., tyrosine^57^, naphthalene^58^, and polyphenol^59^), implicating gut-liver axis in the processing of environmentally derived compounds. These metabolites originate from aromatic substrates that are first transformed by the gut microbiota into intermediate phenolic compounds^60^. After absorption into the portal circulation, these microbial products undergo extensive phase II detoxification in the liver, particularly sulfation, yielding their sulfate-conjugated forms^61^. The levels of these sulfated metabolites in circulation therefore reflect not only the extent of environmental exposures, such as smoking, but also the combined metabolic activities of gut microbial enzymes and hepatic biotransformation pathways.

The association between stressful life events and metabolites was similar in participants with established diagnoses of depressive and anxiety disorders and healthy controls without psychiatric disorders. In the present findings, no significant interactions of stressful life events with psychopathology status on metabolite levels were observed, which may reflect the limited statistical power due to small sample size. A recent NESDA study exploring the metabolomics profile of anxiety disorders identified 67 metabolites related to current anxiety disorder and its subtypes (unpublished work)^25^. Of them, 14 metabolites overlapped with those related to stressful life events, such as two bile acids (i.e., glycoursodeoxycholate and taurodeoxycholate). In parallel, another NESDA study identified 139 metabolites associated with current MDD, with negatively associated metabolites similarly enriched in the long-chain MUFAs^23^. Among these, 29 metabolites overlapped with the stressful life events-related metabolites identified in the current study (see Supplementary Table 11). Notably, several fatty acids, particularly the medium-chain fatty acids such as 10-undecenoate (11:1n1)), showed lower levels in participants with stressful life events and a similar pattern could be found in those with MDD. These fatty acids are typically involved in mitochondrial fatty acid oxidation processes including β-oxidation. Previous studies suggested that under conditions characterized by active fatty acid oxidation (e.g., fasting or high-fat diet), higher levels of carnitines with medium-chain fatty acids could be observed^62^. In contrast, the lower levels of these fatty acids in individuals with stressful life events or MDD may indicate altered mobilization of lipids and potentially decreased activity of lipid oxidation. Taken together, findings from previous studies and the present one suggest that stressful life events, depression and anxiety may cumulatively lead to metabolic alterations by partially converging on the same biological pathways.

The current study had some strengths. To our knowledge, this is the largest study conducting untargeted metabolomics analysis on stressful life events, additionally including external replication in two independent cohorts from populations of different cultural and ancestral backgrounds. However, some limitations should be noted. First, the relatively small sample sizes of the replication cohorts along with differences in lifestyle, cultural contexts and measurements of stressful life events and metabolites in the GBCS might limit the statistical power, leading to the lack of significance for many stressful life events-metabolite associations. Nevertheless, the consistent direction of effect estimates in the NEO study with those observed in NESDA suggested that replication supported the main findings. Future studies are warranted to use the exact same measurements of stressful life events and metabolites in larger sample size to further replicate our findings in more diverse ethnic populations. Second, the cross-sectional design of our study did not support causal inferences. Thus, future studies should examine the significant metabolites identified in a longitudinal design. Third, several relevant life events, such as relocation, long-term caregiving for a seriously ill family member and major workplace conflicts, were not included. This may cause an incomplete assessment of stressful life events, potentially leading to an underestimation of their associations with metabolites. Fourth, some lifestyle factors, such as dietary information, were not collected at the time of metabolic assessment. Given that several metabolites related to stressful life events have been suggested to originate from microbial metabolism of dietary components (e.g., polyphenols), further explorations into the relationship among stressful life events, diet and metabolites are warranted.

## Conclusions

Overall, our study presents the largest comprehensive metabolomic analysis of stressful life events to date. We identified significant associations between stressful life events and specific circulating metabolites, characterized by dysregulation in lipid metabolism, including the bile acid pathway. These metabolic disruptions may potentially indicate stressful life events-triggered changes in lifestyles and gut microbiome. Moreover, the stressful life events-related metabolites showed substantial overlap with those implicated in psychological and cardiometabolic diseases, suggesting that metabolic alterations in the long-chain MUFA and phosphatidylethanolamine might be the potential mechanism linking stressful life events to adverse health outcomes. Therefore, our findings highlight that the metabolomic signature of stressful life events may provide novel insights into the underlying biological mechanisms of stressful life events-related disease risks and inform targeted monitoring to promote health outcomes.

## Data Availability

All data produced in the present study are available upon reasonable request to the authors.

## Acknowledgments

Metabolomics data was generated by the Alzheimer’s Gut Microbiome Project (AGMP) and the Alzheimer’s Disease Metabolomics Consortium (ADMC). The AGMP and ADMC were funded wholly or in part by the following grants and supplements awarded to Rima Kaddurah-Daouk at Duke University in partnership with a large number of academic institutions: R01AG046171, RF1AG051550, RF1AG057452, R01AG059093, U19AG063744, 3U19AG063744-04S1, RF1AG058942, 1U01AG088562, U01AG061359, R01AG081322). As such, the investigators within the AGMP and the ADMC, not listed specifically in this publication’s author’s list, provided analysis-ready data, but did not participate in designing the study, conducting the analyses or writing of this manuscript. A listing of AGMP Investigators can be found at: https://alzheimergut.org/meet-the-team/. A listing of ADMC investigators can be found at: https://sites.duke.edu/adnimetab/team/. Matthias Arnold and Gabi Kastenmüller received funding (through their institutions) from the National Institutes of Health/National Institute on Aging through grants RF1AG058942, RF1AG059093, U01AG061359, U19AG063744, and R01AG069901. Rick Jansen received funding from ZonMw: The Netherlands Organization for Health Research and Development (project number: 636310017, research program GGZ). Brenda Penninx is supported by the research project ‘Stress in Action’ (www.stress-in-action.nl) which is financially supported by the Dutch Research Council and the Dutch Ministry of Education, Culture and Science (NWO gravitation grant number 024.005.010).

The infrastructure for the NESDA study (http://www.nesda.nl) is funded through the Geestkracht program of the Netherlands Organisation for Health Research and Development (ZonMw, grant number 10-000-1002) and financial contributions by participating universities and mental health care organizations (VU University Medical Center, GGZ inGeest, Leiden University Medical Center, Leiden University, GGZ Rivierduinen, University Medical Center Groningen, University of Groningen, Lentis, GGZ Friesland, GGZ Drenthe, Rob Giel Onderzoekscentrum). The NEO study is funded by the participating departments, the Division and the Board of Directors of the Leiden University Medical Centre, and by the Leiden University, Research Profile Area ‘Vascular and Regenerative Medicine’. The GBCS is funded by the University of Hong Kong Foundation for Educational Development and Research (SN/1f/HKUF-DC; C2040028505200), the Health Medical Research Fund (HMRF/13143241) in Hong Kong, the Guangzhou Public Health Bureau (201102A211004011), the Natural Science Foundation of Guangdong (2018A 030313140), Guangzhou Twelfth People’s Hospital, the University of Birmingham, UK, School of Public Health, the University of Hong Kong and Greater Bay Area Public Health Research Collaboration. Metabolomics data of the GBCS was supported by the Natural Science Foundation of Guangdong (2025A1515011659) and the fundamental research funds for the central universities, Sun Yat-Sen university (24qnpy211).

## Authors’ contributions

Yumeng Tian: Writing-original draft, Visualization, Formal analysis. Ruifang Li-Gao: Conceptualization, Writing-original draft, Visualization. Tahani Alshehri: Writing-original draft, Visualization, Methodology. Christopher R. Brydges: Writing-review & editing. Matthias Arnold: Writing-review & editing. Siamak Mahmoudiandehkordi: Writing-review & editing. Gabi Kastenmüller: Writing-review & editing. Dennis O. Mook-Kanamori: Writing-review & editing. Frits R. Rosendaal: Funding acquisition, Supervision, Writing-review & editing, Resources. Erik Giltay: Writing-review & editing. Lin Xu: Funding acquisition, Writing-review & editing. Jiao Wang: Funding acquisition, Writing-review & editing. Rick Jansen: Writing-review & editing. Thomaz Bastiaanssen: Writing-review & editing. Brenda W.J.H. Penninx: Funding acquisition, Writing-review & editing, Resources. Rima Kaddurah-Daouk: Funding acquisition, Writing-review & editing, Resources. Yuri Milaneschi: Conceptualization, Supervision, Writing -review & editing.

