## Supplementary information for "The Metabolomic Signature of Stressful Life Events"

**Supplementary Appendix**

**Study design of NESDA, NEO study and GBCS**

The NESDA is an ongoing longitudinal cohort study on the course and consequences of depression^1^. Briefly, at baseline 2,329 persons with a remitted or current Diagnostic and Statistical Manual of Mental Disorders Fourth Edition (DSM-IV) based depressive (major depressive disorder, dysthymia) and/or anxiety disorder (panic disorder, social phobia, agoraphobia, generalized anxiety disorder), 367 of their siblings and 652 healthy controls, yielding a total of 3,348 participants were included from the community, primary care, and secondary care settings between 2003 and 2007. Follow-up visits were taken place after 1, 2, 4, 6, and 9 years. Each assessment included a diagnostic interview to assess the presence of depressive and anxiety disorders, a medical examination, and several questionnaires on symptom severity, other clinical characteristics and lifestyle. Fasting blood was collected at baseline and the 6-year follow-up. The study was approved by the ethical review boards of participating centers, and all participants provided informed consent. Totally, 2,770 participants (2,463 from baseline and 307 siblings recruited at the 9-year follow-up) and 1,805 participants of the 6-year follow-up (of which 1,685 overlapped with baseline) were measured with Metabolon metabolomics platform will be used as discovery cohort^2^.

The NEO study is a population-based cohort study of individuals aged between 45 and 65 years with a self-reported body mass index (BMI) of 27 kg/m^2^ or higher, living in the greater area of Leiden (in the West of the Netherlands)^3^. In addition, all inhabitants aged between 45 and 65 years from one municipality (Leiderdorp) were invited, irrespective of their BMI. Recruitment of participants started in September 2008 and completed at the end of September 2012. The study was approved by the medical ethical committee of the Leiden University Medical Centre. Participants were invited to come to the NEO study center of the LUMC for one baseline study visit after an overnight fast. The participants were asked to bring all medication they were using in the month preceding the study visit to the NEO study site, both prescribed medication as well as self-medication, such as vitamin supplements. A blood sample of 108 mL was taken from the participants after an overnight fast of at least 10 hours. A subset of participants (N=599) with fasting blood using the Metabolon metabolomics platform will be utilized for external replication analyses^4^.

The GBCS is a four-way collaboration among Guangzhou Twelfth People’s Hospital and the Universities of Hong Kong, Birmingham and Sun Yat-Sen^5^. Participants were drawn from the Guangzhou Health and Happiness Association for the Respectable Elders (GHHARE), from September 2003 to January 2008. The GHHARE was unofficially aligned with the municipal government and had branches in all districts of Guangzhou, the capital city of Guangdong Province in Southern China. About 7% of Guangzhou residents in this age group were included in the GHHARE. The Guangzhou Medical Ethics Committee of the Chinese Medical Association approved the GBCS and all participants gave written, informed consent before participation. All participants were invited to return for the first (March 2008 to December 2012) and second follow-up examinations (March 2013 to January 2020). Both the baseline and follow-up examinations included a face-to-face, computer-assisted interview conducted by trained nurses to collect information on demographic characteristics, lifestyle factors, and family and personal medical history. Anthropometric and clinical parameters were measured. The follow-up questionnaire and clinical and laboratory examinations were largely similar to those conducted at baseline. The reliability of the questionnaire was tested by randomly recalling 200 participants for re-interview and the results were satisfactory. A subset of participants (N=200) with fasting blood using Level One 500 metabolomics platform will be utilized for external replication analyses^6^.

**Measurements of metabolomics and quality control in NESDA**

Metabolomic profiles were measured at baseline, 6-year follow-up and in the sibling sub-study using Metabolon^TM^ Discovery HD4 platform (Metabolon Inc., Durham, North Carolina, USA)^1^. The quality control of metabolites has been reported in the previous NESDA studies^2,7^. Briefly, plasma samples were stored at -80 °C and sent for analyses in two batches. Plasma samples were divided into four fractions; two for ultra-high performance liquid chromatography-tandem mass spectrometry (UPLC-MS/MS; positive ionization), one for UPLC-MS/MS (negative ionization), and one for a UPLC-MS/MS polar platform (negative ionization). Peaks were quantified using the area-under-the-curve in the spectra. To account for run-day variations, peak abundances were normalized by their respective run-day medians. Compounds were identified using an internal spectral database as previously used in various large metabolomic analyses of other samples^8,9^. Pathway enrichment analysis was conducted by using pathways preassigned to the metabolites by Metabolon. In addition, compounds here referred to as partially characterized molecules, have an undetermined chemical identity. They consist of recurring biological entities detected across various studies completed at Metabolon, enabling their identification as unique metabolites despite the lack of full structural elucidation^8^.

Reference sample was used to control for technical measurement variability. Samples with high missingness (>5 SD + mean missingness) were excluded from the dataset. We also excluded metabolites missing across ≥30% of all samples. If outliers or measurement issues were observed, all values on that plate were set to ‘NA’. All samples were normalized to the batch median and metabolites with a technical measurement variability >30% were excluded. Missing metabolite values were imputed with the k-nearest neighbor approach (k=10) since missingness across the remaining metabolites did not accumulate in one measurement wave. The final dataset was log2 transformed and for each metabolite outliers were Winsorized to 5SD away from the mean, and the obtained values were scaled to SD units for comparison. Finally, 820 metabolites, including 681 characterized and 139 uncharacterized, were measured in the NESDA^2,7^.

**Measurements of metabolomics and quality control in NEO study and GBCS**

Metabolomic profiles were measured at baseline using the Metabolon^TM^ Discovery HD4 platform in the NEO study^4^. The quality control has been reported in the previous NEO study, adopting a specific imputation workflow for missing value with multivariate imputation by chained equations (MICE) approach of metabolomic data^10^. Briefly, for the endogenous and unannotated metabolites we applied the multiple imputation method along with a select number of correlated metabolites as auxiliary variables to generate the imputed dataset. Xenobiotic metabolites were imputed to zero to account for true missingness. Then, the dataset was log2 transformed, and values were scaled to SD units for comparison. In the GBCS, metabolomic profiles were at the first follow-up using the untargeted liquid chromatography-mass spectrometry (LC-MS)-based metabolomics platform (Level One 500)^6^. This platform could detect more than 2,000 metabolites in accordance with the Metabolomics Standards Initiative and report relative (semi-quantitative) metabolite abundances^11^, which classified metabolites into four levels, including identified compounds (level 1), putatively annotated compounds (level 2), putatively characterized compound classes (level 3), and unknown compounds (level 4). Metabolite values below the limit of detection were imputed with half of the minimal values. Then, the dataset was log2 transformed, and values were scaled to SD units for comparison. Although the measurements of stressful life events and metabolites in the GBCS differed from those applied in NESDA, we aimed to replicate the findings in a Chinese population to identify consistent associations across diverse ancestral and cultural contexts, lifestyles, and disease profiles.

**Covariates**

Sex, age, level of education, shipment batch, and wave are included as covariates in the main analysis (model 1). In an additional model conducted exclusively in the NESDA, we will examine the impact of further adjustment for lifestyle and health-related factors that may potentially represents mechanisms explaining the associations detected, such as BMI, alcohol consumption, smoking, level of total physical activity, the number of 20 chronic diseases, and medication use (model 2). Specifically, sex (men/women) and education level (basic/intermediate/high) consisted of time-invariant covariates, while age (continuous), BMI (continuous), smoking (never/former/current), alcohol consumption (continuous), and total physical activity level (continuous), self-reported chronic diseases score (based on 20 conditions) (continuous), and lipid-lowering medication use (no/yes) were entered as time-varying covariates. BMI is calculated as measured weight divided by height squared. Alcohol consumption is measured as units per week and smoking status is classified into current, ex- and never-smokers. Total physical activity is assessed using the International Physical Activity Questionnaire (IPAQ), with the total energy expenditure expressed in metabolic equivalent total (MET) minutes per week^12^. self-reported chronic diseases score is defined by the number of self-reported 20 somatic diseases for which one received treatment^13^. Medication use is assessed through medication container inspection of all medications that participants used in the past month. An earlier drug-metabolite study identified three class of commonly prescribed drugs related to widespread metabolite alterations: lipid lowering, anti-hypertensive and anti-diabetics medications^14^. However, while the association with anti-diabetics was mainly driven by the disease of indication, the association with anti-hypertensive medications was mainly driven by co-medication, particularly statins. Thus, we only consider the use of lipid-lowering drugs in the present study.

**List of Threatening Experiences Questionnaire**

| **List of Threatening Experiences Questionnaire** | **Answer: Yes/No** |
| --- | --- |
| You were seriously ill, wounded, or victim of violence |  |
| A close relative was seriously ill, wounded, or victim of violence |  |
| A parent, child, brother, sister, or partner died |  |
| A good friend or close relative died |  |
| You and your partner separated |  |
| You ended a longstanding relationship with a friend or relative |  |
| You had a serious problem with a close friend, relative, or neighbor |  |
| You became unemployed or looked for a job without result |  |
| You were fired |  |
| You were facing serious financial problems |  |
| You had contact with the police or court by misdemeanor |  |
| Money or something valuable was stolen or lost |  |

**Ten items of stressful life events in the GBCS in China**

| **Ten items of stressful life events** | **Answer: Yes/No** |
| --- | --- |
| You were victim of violence |  |
| You experienced a major injury or traffic accident |  |
| Your spouse died |  |
| A close relative had a major illness or passed away |  |
| You experienced a major natural disaster (i.e., flood and drought) |  |
| You and your partner separated or divorced |  |
| You lost your job or retired |  |
| Your business went bankrupt |  |
| You had major conflict within your family |  |
| You lost income or were living on debt |  |
